# Comparing the Fit of N95, KN95, Surgical, and Cloth Face Masks and Assessing the Accuracy of Fit Checking

**DOI:** 10.1101/2020.08.17.20176735

**Authors:** Eugenia O’Kelly, Anmol Arora, Sophia Pirog, James Ward, P. John Clarkson

## Abstract

**Introduction:** The COVID-19 pandemic has made well-fitting face masks a critical piece of protective equipment for healthcare workers and civilians. While the importance of wearing face masks has been acknowledged, there remains a lack of understanding about the role of good fit in rendering protective equipment useful. In addition, supply chain constraints have caused some organizations to abandon traditional quantitative or qualitative fit testing, and instead, have implemented subjective fit checking. Our study seeks to quantitatively evaluate the level of fit offered by various types of masks, and most importantly, assess the accuracy of implementing fit checks by comparing fit check results to quantitative fit testing results.

**Methods:** Seven participants first evaluated N95 and KN95 masks by performing a fit check. Participants then underwent quantitative fit testing wearing five N95 masks, a KN95 mask, a surgical mask, and fabric masks.

**Results:** N95 masks offered higher degrees of protection than the other categories of masks tested; however, it should be noted that most N95 masks failed to fit the participants adequately. Fit check responses had poor correlation with quantitative fit scores. All non-N95 masks achieved low fit scores.

**Conclusion:** Fit is critical to the level of protection offered by masks. For an N95 mask to provide the promised protection, it must fit the participant. Performing a fit check was an unreliable way of determining fit.

## Introduction

During the course of the COVID-19 pandemic, the importance of face mask fit has become apparent, while testing to ensure fit has decreased (1,2). It is known that respiratory protective equipment is only effective when there is an adequate seal formed between a mask and the person’s face to ensure that inhaled air is actually filtered. Indeed, research has suggested that an ineffective seal is the primary cause of airborne contamination amongst wearers of face mask (3). It has been noted that leakage around the face mask may account for a third of airflow across surgical masks and a sixth of the flow across respirators(3). Fit is recognized as being particularly important when determining whether masks are capable of reducing the spread of fine particles, it is normal practice for respirators to be fit tested before use in clinical practice.

COVID-19 has strained supply chains of masks and fit testing supplies while simultaneously placing heavy workloads on hospital staff. This has led to many healthcare facilities having to abandon normal fit testing procedures(1). Qualitative and quantitative fit testing has been replaced by self-performed fit checks, in which the user feels for air leaks(4). The potential impact of abandoning or replacing fit testing procedures on respiratory safety remains understudied.

For individuals outside of healthcare facilities, the Center for Disease Control (CDC) in the United States and Public Health England (PHE) in the United Kingdom have advised the general public to wear fabric face coverings in public(5,6). Notably, there is little literature exploring the fit of fabric face coverings and how much protection they may offer wearers where fit issues are present. Even in normal times, fit testing is reserved for the use of N95 masks and research into the importance of fit for other mask types, such as surgical masks, is limited.

Our study explores the fit and associated protection offered by a range of face masks types. Notably, we test a pre-COVID-19 manufactured N95 mask; a KN95 mask, a standard surgical mask, and a selection of fabric face coverings. We aim to elucidate: importance of fit for protecting the wearer, how well a simple fit check predicts fit, and the relative degree of protection offered by each mask type.

## Methods

### Masks Tested

Three categories of face masks were tested for fit: five N95 masks from different manufacturers, two KN95 brands, surgical style masks, and a selection of fabric face coverings. The N95 masks tested are listed in Table 1. Each participant completed one quantitative fit test and one subjective fit-check per mask.

**Table 1:** Model number, manufacturer, manufacturer information, country of manufacture, and NIOSH approval numbers for the five N95 masks tested.

| **Model** | **Manufacturer** | **Manufacturer**<br>**Headquarters** | **Country of**<br>**Manufacture** | **NIOSH Approval**<br>**(TC) Number** |
| --- | --- | --- | --- | --- |
| 8511 | 3M | Saint Paul, USA | USA | TC-84A-1299 |
| 8200 | 3M | Saint Paul, USA | USA | TC-84A-4271 |
| AP0028 | Aero Pro Co<br>Ltd | Taipei, Taiwan | Taiwan | TC-84A-4175 |
| 9500 | Makrite | Tianzhong, Taiwan | China | TC-84A-5411 |
| ZYB-11 | Zhong Yi | Wuhan, China | China | TC-84A-7877 |

The KN95 mask was manufactured by Zhong Jian Le KN95 mask manufactured by Chengde Technology Co LTD, China and certified according to Chinese standard GB2626-2006.

Five basic fabric masks were tested on three of the participants. These included a 100% cotton bandana, a mask made of stretch material, a pleated surgical-style mask, and two masks designed to contour to the face. All of these masks were of simple construction, made of one or two layers of fabric and containing neither a filter nor fitting aids such as a nose wire.

N95 and KN95 masks were worn for at least five minutes before testing to purge interior particles. Surgical masks and fabric face coverings, which were non-sealing, were worn for at least three minutes before testing.

### Fit Checks

Participants were asked to don the mask and perform a fit check according to UK National Health Service (NHS) guidelines (7). Participants were asked to feel around the sides of the mask and report if they could feel any air leaks, suggesting poor fit. They were also instructed to feel if the mask responded to heavy breathing, with the mask being pulled more tightly against the face during a sharp inhale. Participants reported the result of the fit check (a belief the mask fit properly, or a belief the mask did not fit properly). They were then asked how confident they were in their assessment, with “high” denoting a high level of confidence that their prediction of mask fit was correct, “medium” denoting a medium level of confidence, and “low” denoting little confidence in their fit check result. To assess the reliability of the fit-checking method, fit check answers were compared with the quantitative fit scores for each mask.

### Quantitative Fit Testing

Quantitative fit testing was used to determine actual mask fit. Quantitative fit testing simultaneously assesses the number of particles inside and outside a mask. By comparing these particle counts, a fit factor score is given. Fit factor scores provide a quantitative measure of the degree of protection wearers might expect. While fit factor scores generated in a testing environment do not exactly predict the degree of workplace protection, fit factors have been shown to be an effective method of predicting actual workplace protection(8)

Qualitative fit testing was performed with a TSI PortaCount Pro Respirator Fit Tester model 8038+, capable of assessing N95 mask fit. The Portacount 8038 measures particles with a minimum size of 0.02 micrometers at a sampling flow rate of 350 cm^3/min. Fit tests were conducted using OSHA protocol 29CFR1910.134 (https://www.osha.gov/laws-regs/regulations/standardnumber/1910/1910.134). A minimum fit score of 100 must be achieved for N95 masks to offer appropriate protection according to OSHA guidelines(9). A fit score of 200+ represents the highest score possible for the masks tested.

Seven participants wore face masks. Three participants were male and four participants female. Participant ages ranged from 17 to 74, with a mean age of 45 and a median age of 51. Participants completed seven activities as defined by the OSHA fit-testing protocol. This range of activities is intended to represent the spectrum of normal occupational activity, and thus reproduce fit during daily activity.

Two modifications were made to the 29CFR1910.134 protocol. First, participants were allowed to sit through sections of the fit testing. Secondly, participants were allowed to adjust their masks if needed, with the researcher making note if adjustments were necessary during the fit test. This allowance was made to allow us to complete testing on poorly fitting masks.

### Calculating Fit Factor

Fit factor was calculated by the PortaCount 8038 using the formula:

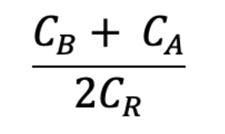

Where Cb is the ambient particle concentration before the respirator sample is taken, Ca is the ambient particle concentration after the mask sample is taken, and Cr is the mask particle concentration. This formula is used to account for normal fluctuations in ambient particle levels.

As the fit factor reflects the number of particles filtered by the masks, it can be considered a numerical score of how much protection the mask provides on any individual wearer. Half face respirators, such as N95 masks, must achieve a fit of at least 100 to be worn as protective gear, according to OSHA guidelines(9). The maximum score achievable on N95 setting of the Portacount 8038 is over 200, which is represented as 200+.

## Results

### Protection and Fit

The 3M model 8511 N95 mask fit the greatest number of participants. Three out of the seven participants achieved a ‘pass’ value while wearing the 3M 8511 mask, two of whom received the maximum fit factor. Four of the participants failed the fit test with the 3M 8511 model.

The Xiantao Zong mask and Aero Pro mask fit none of the participants. The Xiantao Zong mask had fit factors with a mean of 13.2, and a median of 5.6. The Aero Pro had a mean of 35.5 and a median of 21. The 3M 8200 fit two out of the seven participants, with a mean of 72.3 and a median of 64. The Makrite mask fit one out of the seven participants with a mean of 37.7, a median of 14.

The KN95 mask had a visible poor fit and showed very low scores, with an average fit factor of 2.2 Minimal variation was experienced between participants.

The surgical mask showed similar fit scores to the KN95 mask with an average fit score of 3.2 and median fit score of 2.2.

The fabric face masks also showed similar fit scores, with an average of 2.1 and a median score of 2.1.

**Figure 1:**
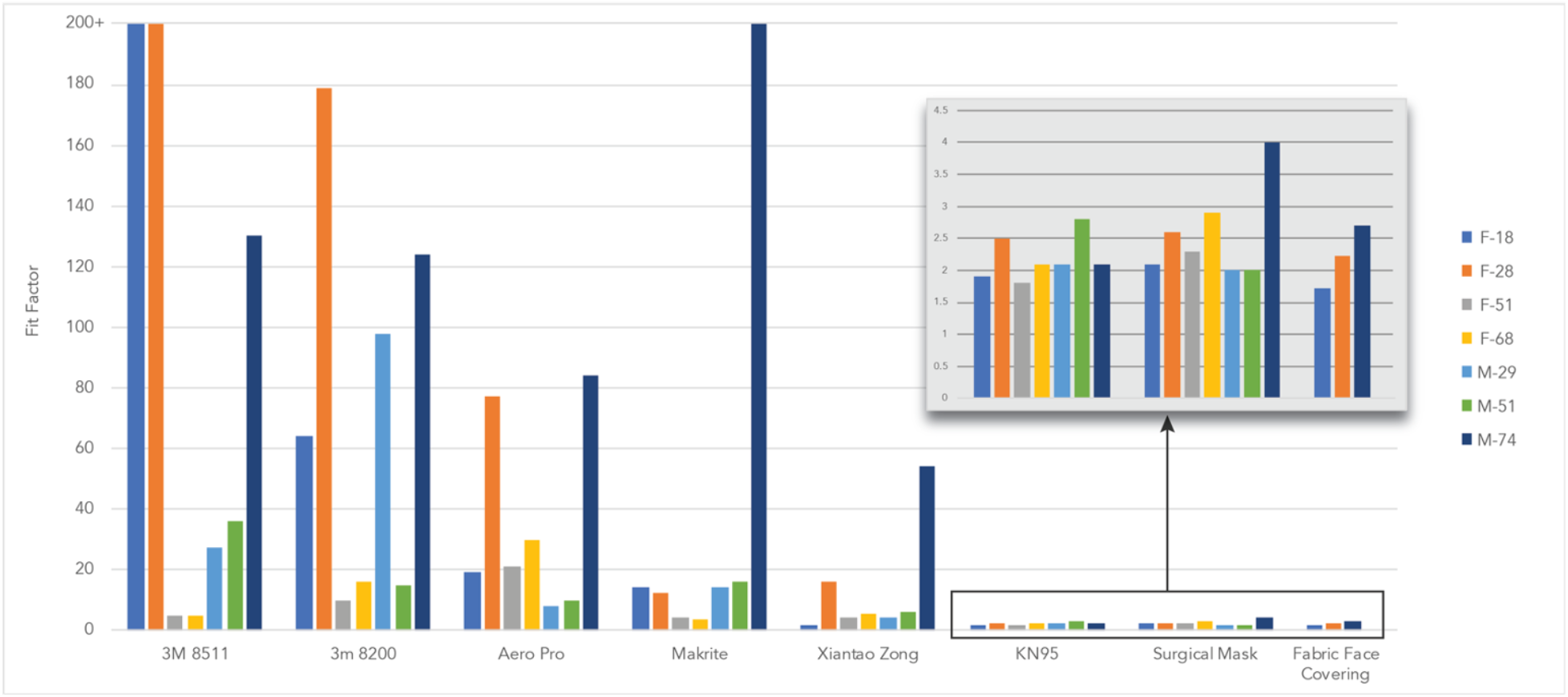
The fit factor achieved by a set of volunteers while wearing different varieties of masks. Protection when wearing an N95 respirator was high only if the respirator properly fit the participant. Fit factors for KN95 masks, surgical masks, and cloth masks were similar. It is possible an error with the testing apparatus created artificially high scores for M-74 while wearing the surgical mask.

While fit factor scores are derived from the number of particles filtered by the mask, the fit factor numbers are not directly equal to filtration efficiency. Measurements of particles counts inside and outside of the mask were compared and are illustrated in figure 2 which represents the number of interior vs. exterior particles in the last cycle of the test.

**Figure 2:**
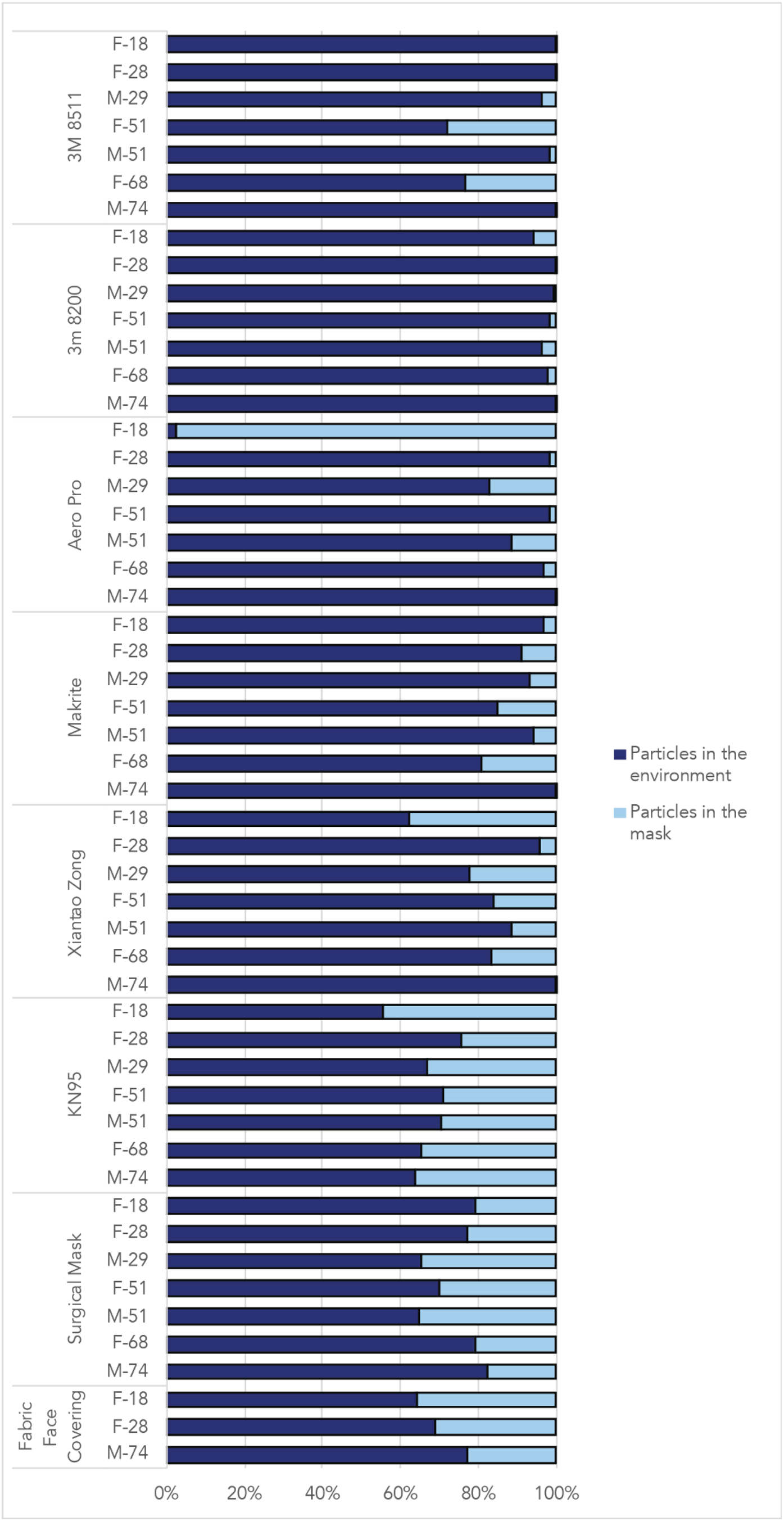
Percentage of particles outside of the mask and percentage of particles measured inside of the mask during the last cycle of normal breathing during the fit test.

### Fit Checks

All participants were able to correctly identify the lack of fit offered by the KN95 mask, likely due to the visibly poor fit by which the mask failed to sit flush to the face.

Out of 35 tests on N95 masks, participants believed 17 masks fit, 2 with low confidence, 7 with medium confidence, and 7 with high confidence. 6 of these masks did indeed fit the wearer, leading to a 35% accuracy rate of predicting a mask fit.

Out of the 35 tests, participants believed 18 masks did not fit, 7 with medium confidence and 11 with high confidence. During all 18 of these tests, the testers themselves correctly identified that the mask did not fit, thus leading to a 100% accuracy when predicting lack of fit.

If fit checks accurately predict fit, it is expected that qualitative fit score would be a function of the fit check results and fit check confidence. By extension, masks which passed the fit check would have higher qualitative fit scores. In fact, this was not the case (Figure 3). The mean fit of masks which were considered to fit with only a low degree of confidence was 138.5. The mean of masks considered to fit with a medium degree of confidence was 74.1 while the mean of those believed to fit with a high confidence was 105.8. Even if only correct fit checks were taken into account, there was no correlation between the degree of confidence of fit and quantitative fit.

**Figure 3:**
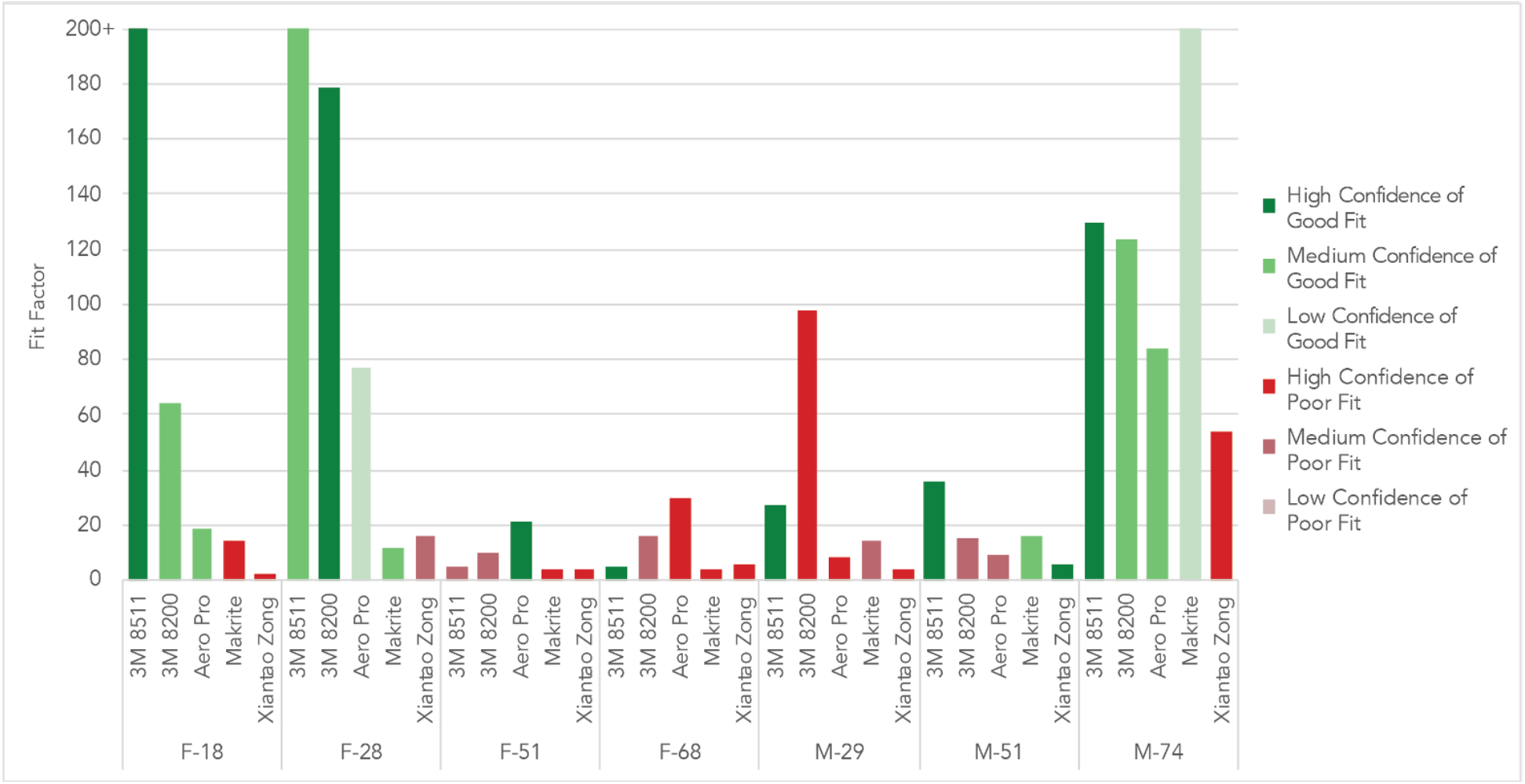
Participants’ fit-check predictions with the qualitative mask fit factor organized by participant. Fit factor results are color coded to represent the participants’ fit check results with green representing a belief of fit and red representing a belief the mask did not fit. Depth of color represented confidence with lighter shades representing low confidence in the fit check results and darker shades representing a high confidence.

When considering masks which did not fit, masks which were believed to not fit with a high degree of confidence should have lower fit factors. Of the masks which were correctly believed not to fit, those believed not to fit with a high degree of confidence had an average score of 18.2 while those believed not to fit with a medium confidence had a lower average of 13.4. Full breakdown of the data can be found in the related open data set.

## Discussion

### Summary of Key Findings

The importance of fit for N95 masks has particular implications during the COVID-19 pandemic as these masks are generally reserved for clinicians at a time hospitals are struggling to cope with demand for conventional fit testing. Proper fit is critical for N95 masks to protect the wearer. Even a small fit issue not detected by the wearer when performing a fit check can greatly decrease the protection offered by the N95 mask. Our results indicate that the method of ‘fit check’, which is being used in many hospitals, is not reliable. The results show that 4 out of 7 participants were unable to achieve a proper fit from any of the tested N95 masks. In addition, all participants made at least one incorrect determination of fit after performing a fit check. These findings suggest that traditional qualitative or quantitative fit testing for individuals in need of respiratory protection is highly advisable. The high proportion of fit test failures, and the associated reduction in effectiveness, has particular implications for healthcare facilities and other work-environments where fit testing may be abandoned during a pandemic.

### Comparing Mask Fit

#### Fit of N95 Masks

These results indicate that proper fit is essential for N95 masks to provide high degrees of protection. N95 masks which fit the wearer tended to filter more than 95% of airborne particles and offer superior protection. Poorly fit N95 masks offered a range of protection, in some cases comparable with surgical and cloth masks.

This study confirms that NIOSH certification that a mask can perform at N95 levels alone is insufficient if the mask is poor fitting. Proper fit is absolutely necessary if the mask is to offer the wearer protection. Furthermore, our results indicate it is not enough to assume that any N95 mask will be likely to fit the majority of a population. The most widely fitting mask, the 8511 N95, fit only three out of the seven participants. Other masks, such as the Aero Pro and Xiantao Zong, did not fit any of the participants adequately.

#### Fit of KN95 Mask

The KN95 mask did not fit the participants. The KN95 masks provided a loose fit with easily visible gaps along the entirety of its connection with the face.

#### Fit of Surgical Masks

Surgical masks are not designed to seal to the face and thus do not provide wearers with the same level of protection as an N95 mask. Nonetheless, our results indicated that surgical masks blocked over 40% of particles for all participants in the last cycle of the quantitative fit test.

#### Fit of Fabric Face Masks / Coverings

The fabric face masks tested produced similar fit factors to commercial KN95 masks and surgical masks; however, the masks used were of the most basic design and construction. It is likely that an improved design coupled with the use of better materials would raise the effectiveness of these masks.

### Implications of Findings

In healthcare settings, clinicians are provided with N95 masks, or equivalent (e.g. FFP3), where procedures are being performed which are associated with a high risk of viral transmission. Existing literature is limited but has noted that poor fit can severely alter the effectiveness of clinical respiratory protective equipment, such as N95 masks. Such research has indicated that better fit is associated with increased protection in controlled testing environments, but it also has been noted that this may not actually result in decreased rates of infection amongst clinicians who engage in fit testing(8). In normal circumstances, the fit of a mask is assessed by a professional fit test before clinical use in hospitals. Importantly, the pandemic has disrupted normal fit testing processes as there is both inadequate range and quantity of masks to satisfy conventional testing requirements(1). There is also a limited range of alternative masks available to facilities in the event that an individual fails a fit test.

KN95 and surgical masks are also used in clinical practice, especially in some countries or when resources are limited such as during this COVID-19 pandemic. These masks are also relatively accessible to the public, though they are not recommended for this purpose due to supply concerns (https://www.mayoclinic.org/diseases-conditions/coronavirus/indepth/coronavirus-mask/art-20485449). In the US, the Food and Drug Administration (FDA) and CDC have both approved the use of KN95 masks where N95 masks are unavailable, even though they are not routinely fit tested. Notably, concerns have been raised about the use of KN95 masks, and in particular, the lack of an adequate seal(10). Whilst the mask itself does have a high filtration capacity, unless a tight seal is achieved, this may be rendered redundant.

It has been suggested that fit testing provides differential benefits for various types of masks, but general principles about face coverings are universal(11). Respiratory protective equipment can only perform at its stated material filtration levels when there is an adequate seal formed between a mask and the person’s face. The space between the wearer’s mouth and the mask acts as an extension of their breathing apparatus. During expiration, breathing movements increase pressure within the apparatus and force air through the filter of the mask. The reverse occurs during inspiration. If an ineffective seal is formed around the mask, contaminated air will take the path of least resistance through gaps around the mask, thus reducing the effectiveness of the mask substantially(12). Our research found that even if a mask is well constructed, you cannot predict the protection it will offer. It is critical to perform fit testing in order to ensure the mask fits properly and is acting as an extension of the breathing apparatus rather than merely a shield to block some of the incoming particles.

### The Reliability of Fit Checks

Participants were unable to reliably predict whether masks fit properly. While no participant mistook a well-fitting mask for a poorly fitting mask, participants routinely believed poorly fitting masks fit them.

Nor did participants’ confidence of a mask’s fit correlate with the actual fit of the mask. Masks believed to fit with a low confidence outperformed masks which were believed to fit with a medium or high degree of confidence. Masks which were believed with high confidence to have a poor fit performed on average better than those believed not to fit with only a medium degree of confidence.

These results suggest that individuals may not be able to accurately assess the fit of their mask through self assessments, such as ‘fit checks’. Three out of the seven participants worked in a healthcare or healthcare-related field and had received a degree of mask fit education. One participant worked in a hazardous industry and had been required to wear face masks or respirators for certain working conditions. These participants were somewhat more likely to correctly identify mask fit through a fit check. All but one of these more experienced participants incorrectly identified 4 out of 5 N95 masks. While this indicates education and experience may be of benefit, a 20% fail rate is still concerningly high.

### Strengths and Limitations

Due to the lengthy nature of qualitative fit testing and the age of some participants, most participants underwent the tests while seated. We do not expect this to have had an impact on the testing results as all NIOSH qualitative fit testing activities were followed except the change from standing to seated. Two participants, M-51 and M-29, had some degree of facial stubble or hair which may have affected fit and the ability to sense air movement.

Participants successfully represented a range of ages and prior experience wearing masks. Three participants had experience working in healthcare related fields, one participant had experience in an industry which can require the wearing of respirators and the remaining three participants did not have any relevant prior experience. Our results were generally concordant within our sample population; however, further studies with larger sample sizes would be necessary to offer definitive conclusions.

## Conclusion

To offer adequate respiratory protection for the wearer, a face mask must not only be made of high filtration, low resistance material, but must also fit the wearer. When a face mask has poor fit, the value of high filtration material decreases such that the wearer achieves similar protection from a fabric face covering as from a poorly fit KN95 mask. Indeed, our results indicated that surgical masks, poorly fit KN95 masks, and basic fabric face coverings offered similar levels of protection to the wearer. These findings suggest that during pandemics or other times when N95 masks are in short supply, it is essential to ensure adequate fit for the wearer. Having a wide variety of mask models and sizes stockpiled is critical as one mask model cannot be assumed to protect the majority of wearers.

## Data Availability

The data set relevant to this study is available in the Cambridge open access data repository. The DOI for this is: https://doi.org/10.17863/CAM.56361

## Acknowledgements

We would like to thank our participants who patiently sat through rounds of qualitative fit testing. Without their willingness to take time out of their busy lives and suffer through wearing uncomfortable masks, this study would not have been possible.

